# Linking mpox wastewater surveillance with reported clinical cases in three countries in Sub-Saharan Africa

**DOI:** 10.64898/2026.06.18.26355929

**Authors:** Elena Sgarabotto, Ananda Tiwari, Melissa Kabena, Eric Lyimo, Palpouguini Lompo, Dylan Shea, Evodie Ngelesi, Jackson P. Mushumbusi, Garba Zakaria, Edward Msoma, Berenger Kabore, Steven C. Mnyawonga, Sibidou Yougbare, Modest Chuwa, Thanh Tam Tran, Elisa Salmivirta, Taru Miller, Annastiina Rytkönen, Rolf Lood, Adriana Krolicka, Marc Christian Tahita, Vito Baraka, Vivi Maketa, Tarja Pitkänen

## Abstract

The emergence of the novel monkeypox virus (MPXV) clade Ib in the Democratic Republic of the Congo (DRC) and neighboring countries in late 2023 highlighted the need for rapid, scalable surveillance approaches to support outbreak detection and response. As part of the ODIN-Mpox project, wastewater surveillance (WWS) systems were established as an emergency public health measure in three Sub-Saharan African countries (DRC, Tanzania, and Burkina Faso) to evaluate the feasibility of wastewater-based monitoring for mpox and strengthen local surveillance capacity. Between January 2025 and April 2026, 117 wastewater samples were collected from selected sites and analyzed for MPXV DNA using targeted qPCR assays. Clinical mpox data were obtained from national surveillance systems and WHO reports to assess epidemiological linkages between wastewater detections and reported infections. Six wastewater samples tested positive for MPXV DNA. During the study period, DRC experienced the highest disease burden, with weekly reported cases peaking at about 3,000 in January 2025, while Tanzania reported a peak of 20 weekly cases in March 2025. No confirmed clinical cases were reported in Burkina Faso. No clear relationship was observed between reported case numbers and qPCR Ct values in positive wastewater samples. Despite the low detection frequency, the project demonstrated the operational feasibility of implementing MPXV wastewater surveillance in resource-limited settings and established laboratory capacity for environmental monitoring of emerging infectious diseases. Given the early stage of WWS implementation in the region, the study identified opportunities for further system strengthening, including optimization of sample processing and reporting workflows, improved access to laboratory supplies, and enhanced integration of environmental and clinical surveillance data streams. These findings highlight the value of WWS as a complementary component of integrated public health surveillance systems and emphasize the need for continued investment in laboratory capacity, harmonized methodologies, governance frameworks, and knowledge exchange to enhance outbreak preparedness and response in low-resource settings.

## 1. Introduction

Mpox outbreak caused by the monkeypox virus (MPXV), has re-emerged as a major public health concern following the rapid spread of the emerging subclades IIb and Ib. The global clade IIb outbreak during 2022–2024 was followed by the emergence of clade Ib in late 2023 in the Democratic Republic of the Congo (DRC) and neighboring countries [1,2]. In comparison to the clade Ia, the clade Ib showed increased transmissibility, which has raised the public health concern [2,3]. The spread of emerging MPXV sub-clades prompted the World Health Organization (WHO) to declare a Public Health Emergency of International Concern (PHEIC) in 2022 due to clade IIb and again in 2024 due to clade Ib [4–6]. Following the WHO PHEIC declarations, the Africa Centers for Disease Control and Prevention (Africa CDC) declared a Public Health Emergency of Continental Security, followed by the DRC Ministry of Public Health declaring a Public Health Emergency of National Concern [7–9].

The global spread of the mpox outbreak, characterized by sustained human-to-human transmission across multiple regions, highlighted the need for rapid, scalable and sustainable surveillance systems capable of supporting outbreak detection and informing public health responses [9,10]. Mpox surveillance currently relies primarily on case-based clinical reporting, in which suspected and laboratory confirmed cases are aggregated and reported over time [11]. Although essential for clinical treatment and case management, clinical surveillance is inherently delayed because it depends on symptom onset, healthcare seeking behaviors, diagnostic testing and reporting, a series of actions that can take days to weeks and may limit the timeliness of outbreak detection [12,13]. Infected individuals shed MPXV in lesion material and multiple body fluids, including saliva, respiratory secretions, urine, and feces, which can enter wastewater systems through bathing, urination and defecation [14–17].

Accordingly, multiple studies have detected MPXV DNA in wastewater during community outbreaks [18–20]. Most published mpox wastewater surveillance (WWS) to date have been conducted in high-income countries, where wastewater infrastructure and laboratory capacity are well established [19–22]. Thus, WWS has increasingly been recognized as a valuable complementary tool for population level infectious disease monitoring [21,23]. Because WWS captures pathogen genetic material shed by infected individuals regardless of healthcare seeking behavior, it can provide insights into community level transmission independent of diagnostic testing coverage [21,23]. During the COVID-19 pandemic WWS demonstrated its value as an early warning system and accelerated interest in applying similar approaches to other pathogens, including MPXV [20,21,23]. However, implementation remains limited in lower- and middle-income countries (LMICs), particularly Sub-Saharan Africa (SSA), due to restricted sewerage coverage, limited laboratory capacity, competing public health priorities, and limited policy uptake [24]. Despite these challenges, WWS may be particularly valuable in LMICs where clinical testing capacity is limited or overwhelmed, as it provides population-level information without requiring individual testing [25].

As part of the ODIN-Mpox project (https://www.global-health-edctp3.europa.eu/projects/odin-mpox_en) [26], this study aimed to establish a wastewater-based environmental surveillance system in three SSA countries: the DRC, Tanzania and Burkina Faso, for the detection and monitoring of mpox using qPCR-based analysis of wastewater. In parallel, the project assessed the relationship between MPXV DNA prevalence detected in wastewater and clinically reported mpox cases in the corresponding catchment areas, to evaluate the performance of WWS in relation to established clinical surveillance systems and its potential as a complementary public health tool. This integrated approach, combining WWS, clinical data, and genomic surveillance, was designed to support both the immediate response to the ongoing public health emergency associated with emerging MPXV subclades and to strengthen longer-term preparedness for future zoonotic threats. In addition, an important objective of the project was to enhance laboratory capacity and foster sustainable collaboration between African and European partners.

## 2. Materials and Methods

### 2.1 Study sites

This study reviewed the sampling locations of the ODIN project (https://cordis.europa.eu/project/id/101103253.) [27], an earlier project of the consortium in the same countries and selected a subset of these sites for WWS of mpox in the present study. Where feasible, priority was given to sampling locations in capital cities, airports and the municipalities with ongoing mpox outbreaks. Wastewater samples were collected from 12 sampling sites across the three SSA countries: the DRC (4 sites), Tanzania (4 sites), Burkina Faso (4 sites) (Figure 1). These sites mainly comprised open wastewater channels draining densely populated urban areas and major transport hubs, enabling the capture of sewage contributions from both resident and transient populations, and also included surface water sites such as docking points for small passenger boats between cities and local bathing areas used by travelers. Most selected sites have a long history of poliovirus WWS, providing established sampling logistics and local familiarity with environmental monitoring practices.

**Figure 1.**
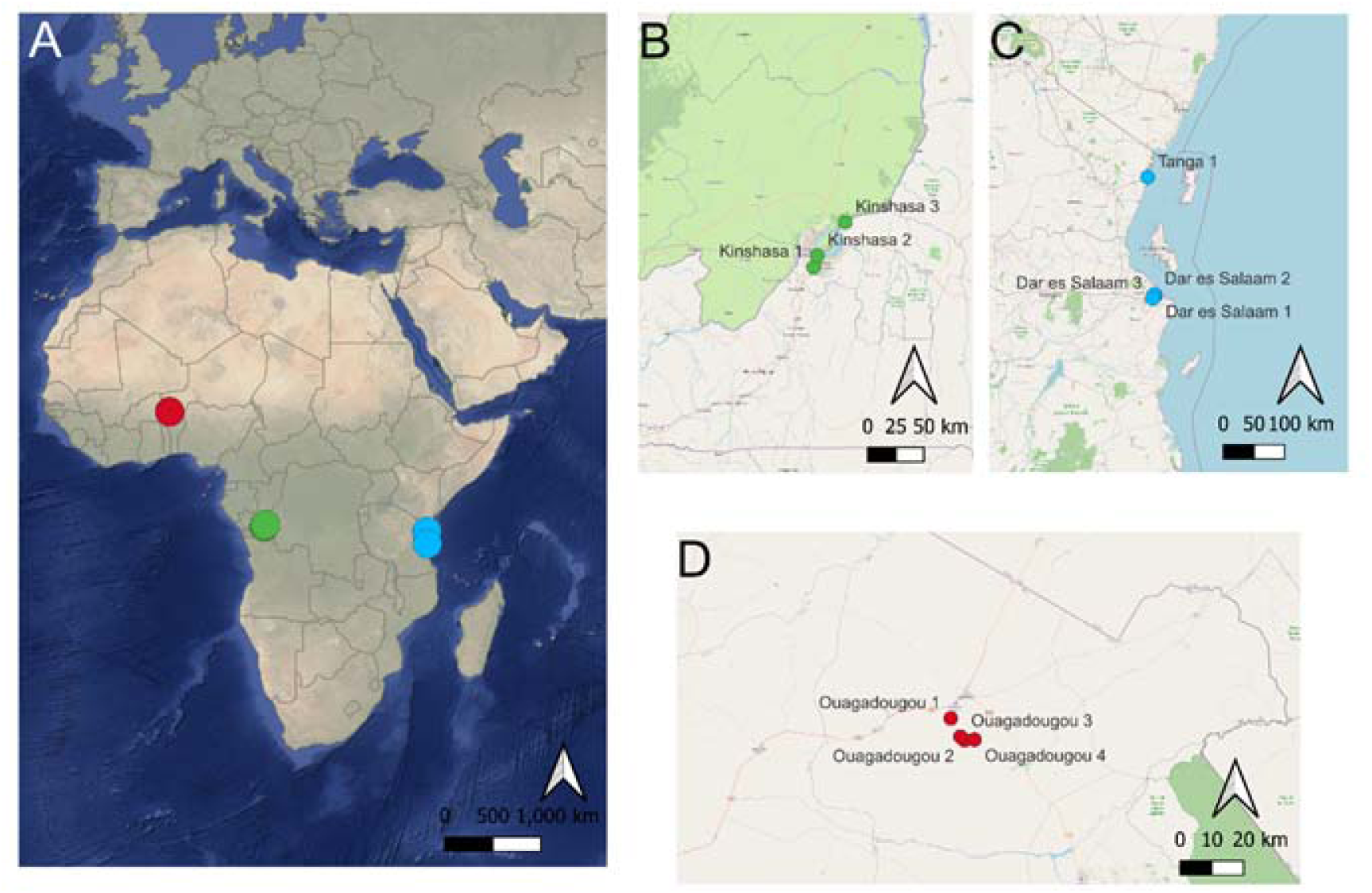
Geographic distribution of wastewater passive sampling sites monitored within the ODIN-mpox project (2025-2026). (A) Continental overview; (B) Democratic Republic of the Congo; (C) Tanzania; (D) Burkina Faso. Map created with QGIS (v 3.36.3).

### 2.2 Harmonization of laboratory workflow

Prior to sample collection and analysis, laboratory workflows were harmonized among the three countries. The ODIN project handbook [28] and standard operating procedures (SOPs) were adapted to local settings, and laboratory molecular workflows were audited. Online and on-site training for sampling and molecular analysis including qPCR was conducted, and an amendment to the ODIN Handbook was circulated among the project partners. Sampling and analytical methods were further evaluated and validated in a single-laboratory study. Additional field training was conducted to ensure harmonized procedures across participating laboratories, including visual guidance for sampler assembly, deployment, and retrieval [29].

### 2.3 Wastewater sample collection

Wastewater was sampled using torpedo passive samplers, deployed in the sewage for 24 to 72 hours to enable continuous accumulation of viral material present in the water [28,30]. This approach provided a robust, low-maintenance method suitable for challenging field conditions and supported consistent sampling across countries. The passive samplers contained three nitrocellulose membrane filters for viral particle capture, as well as absorbent padding for collection of liquid for microbiological analysis [30].

### 2.4 DNA extraction and MPXV DNA detection

After retrieval of the torpedo sampler, two filters from each torpedo were selected for molecular analysis. Nucleic acids were extracted from the filters using the QIAGEN QIAamp Viral DNA Mini Kit (QIAGEN, Venlo, Netherlands) following the manufacturer’s guidelines apart from a modification of the lysis step. Each filter was lysed separately, and the resulting lysates were subsequently combined and purified in a single extraction. Nucleic acid quantification was performed with the Qubit Instrument and the dsDNA quantification kit (ThermoFisher, Waltham, USA). Quantitative PCR (qPCR) was used to detect MPXV DNA. The assay panel included a non-variola Orthopoxvirus generic assay targeting the E9L gene, a generic MPXV assay targeting the G2R_G region, and clade-specific assays designed to differentiate circulating MPXV lineages, including C3L for the clade I Congo Basin lineage, Mpox-K for the clade Ib Kamituga variant, and G2R_WA for the clade II West African lineage (Table 1) [31–33]. When the laboratory readiness and the qPCR instrument allowed, a triplex assay was used to enable simultaneous amplification of three targets (G2G_R, Mpox-K and G2R_WA) within a single reaction, improving throughput and reducing reagent usage whilst being able to maintain target discrimination through fluorophore separation. Instead, singleplex assays for all five targets were used in laboratories whose qPCR instruments did not support multiple fluorescent detection. Both the multiplex and the singleplex qPCR used the SsoAdvanced Universal Probes Supermix chemistry, and reaction followed manufacturer’s instructions (Biorad, Hercules, USA). In Tanzania, a third protocol, the FlexStar Monkeypox virus PCR Detection Mix 1.5 (altona Diagnostic, Plain City, USA), a generic MPXV DNA detection method used in clinical settings was also available for use. The availability of three protocols ensured that all partners could analyze the same targets regardless of instrument capability.

**Table 1.** Primer and probes used for the detection of mpox virus clades and generic non-variola orthopoxvirus targets by singleplex and multiplex qPCR assays.

| Assay | Oligo name | Reference | Assay |
| --- | --- | --- | --- |
| Non-Variola Orthopox Generic (E9L) | E9L.F | Li et al. 2006 [31] | Singleplex |
|  | E9L.R |  |  |
|  | E9L.P |  |  |
| MPOX generic clade I and II (G2R_G) | G2R_G.F | Li et al. 2010 [32] | Singleplex and multiplex |
|  | G2R_G.R |  |  |
|  | G2R_G.P |  |  |
| MPOX Congo Basin specific clade I (C3L) | C3L.F | Li et al. 2010 [32] | Singleplex |
|  | C3L.R |  |  |
|  | C3L.P |  |  |
| Mpox virus clade I b Kamituga variant (Mpox-K) | MpoxK.F | Schuele et al. 2024 [33] | Singleplex and multiplex |
|  | MpoxK.R |  |  |
|  | MpoxK.P |  |  |
| MPOX West African specific clade II (G2R_WA) | G2R_WA.F | Li et al. 2010 [32] | Singleplex and multiplex |
|  | G2R_WA.R |  |  |
|  | G2R_WA.P |  |  |

The performance of the method was verified by using inactivated viral particles of three clades of MPXV (clades Ia, Ib, IIb) and purified nucleic acids were received from WHO Biohub for non-commercial public health use. Representativeness of the wastewater sampling and the efficiency of the DNA extraction were assessed using two internal markers. Fecal indicator bacteria *Escherichia coli* (*E. coli*) enumerated from the liquid collected into the sampler padding served as a control confirming that the torpedo sampler had been in contact with wastewater during the sampling event [34]. A viral marker, CrAssphage, was analysed from the DNA extracted from the filter material and acted as a process control to verify DNA recovery and PCR amplification performance [35]. Furthermore, the CrAssphage assay was used as a human fecal contamination marker in environmental samples together with *E. coli* to assess the levels of human fecal pollution [36,37].

### 2.5 Reported clinical cases

When available, clinical case data from the catchment areas of the WWS sites were obtained from national or regional level epidemiological authorities. In the DRC, weekly mpox case data were acquired from the Ministry of Public Health (Epidemiological Surveillance Directorate), at municipal, regional and national levels [38,39]. In Tanzania, monthly mpox case data were extracted from WHO reports at the national level [38]. For the Tanga WWS site, weekly mpox case data were obtained from the Tanga Regional Epidemiological Surveillance Authority. No confirmed mpox cases were reported in Burkina Faso during the study period [38]. Data was analyzed and visualized with R, package ggplot (v 4.0.0).

## 3. Results

Wastewater surveillance implemented across the participating SSA countries in the ODIN-Mpox project generated a diverse set of samples and MPXV detection results. Variability in surveillance outcomes reflected differences in site coverage and the underlying epidemiological context in each country (Table 2) [40,41].

**Table 2.** Wastewater sampling sites, total number of samples, MPXV-positive wastewater samples, and corresponding confirmed mpox cases in the Democratic Republic of the Congo, Tanzania, and Burkina Faso, January 2025–April 2026. NA, not applicable.

| Country | Number of locations | Number of wastewater samples collected | Number of wastewater samples detected positive | Number of confirmed human cases (Municipality) | Number of confirmed human cases (Country) |
| --- | --- | --- | --- | --- | --- |
| Democratic Republic of the Congo | 4 | 48 | 0 | 94 | 102 573 |
| Tanzania | 4 | 28 | 6 | *52 | 304 |
| Burkina Faso | 4 | 41 | na | 0 | 0 |

\*Note: WWS in Tanzania was conducted in the Dar es Salaam and Tanga regions. Clinical mpox case data were available only for the Tanga region from January to October 2025; no clinical case data were available specifically for Dar es Salaam from the study period, and for Tanga from November 2025 onwards.

### 3.1 Democratic Republic of the Congo

In DRC, a total of 48 wastewater samples were collected across the four sampling locations, between April 2025 and March 2026. Fecal indicator bacteria (*E. coli*) were detected in 4% of samples. Nucleic acids were extracted with mean concentration of 4.68 <u>ng</u>/µL (range 0.20-30.6 <u>n</u>g/µL, median 2.91 <u>ng</u>/µL). MPXV DNA was analyzed using singleplex qPCR assays, with no detections in any samples. CrAssphage was detected in 20% of samples.

At the national level, reported clinical mpox cases reached several thousand per month, with a peak weekly total of approximately 3,000 cases in January 2025 (Figure 2). In contrast, within the municipalities corresponding to the wastewater sampling locations, reported cases remained low, generally below 10 cases per month over the January 2025–March 2026 period (Figure 3).

**Figure 2.**
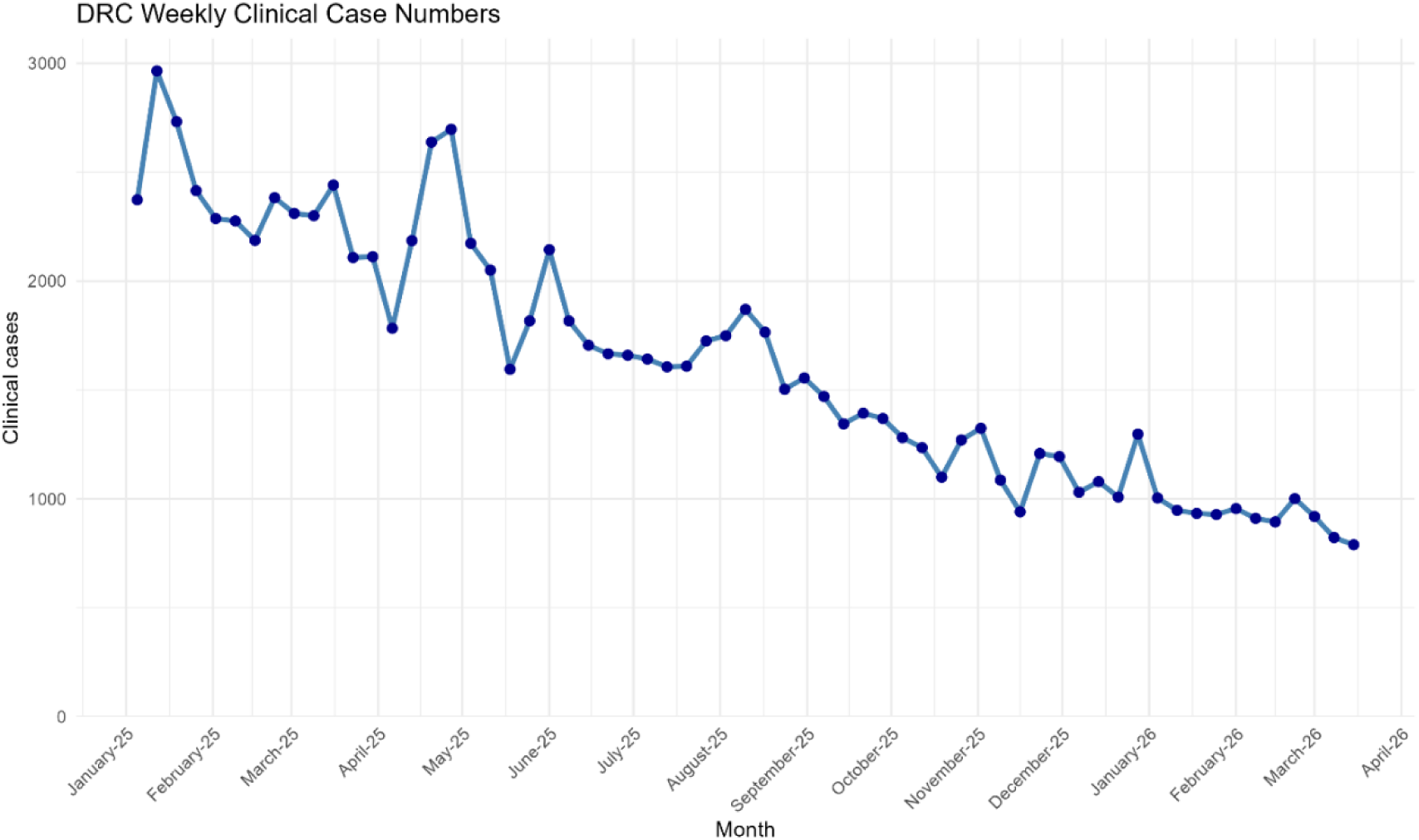
Weekly reported mpox cases in the Democratic Republic of the Congo, January 2025–March 2026 (non–clade specific). Each point represents national weekly case counts. Source: Ministry of Publi Health, Epidemiological Surveillance Directorate (DHIS2 reporting system).

**Figure 3.**
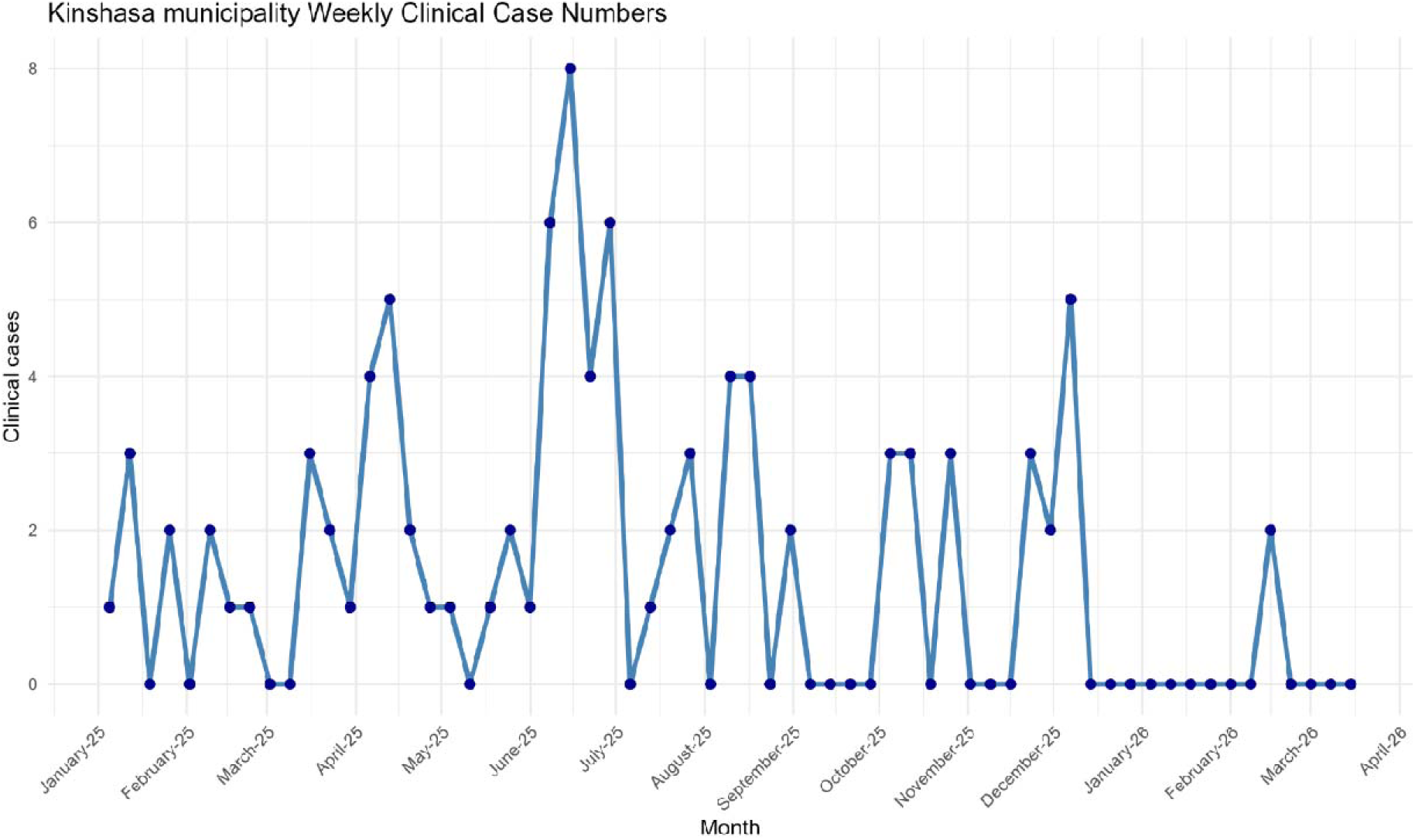
Weekly reported mpox cases in Kinshasa municipalities, January 2025–March 2026. Source: Ministry of Public Health, Epidemiological Surveillance Directorate (DHIS2 system).

### 3.2 Tanzania

A total of 28 passive wastewater samples were collected from four sampling locations between January 2025 to December 2025. *E coli* detection confirmed the presence of fecal material in all samples. Nucleic acids were successfully extracted from 17 samples, with a mean concentration of 7.61 <u>n</u>g/µL (range: 2.63–17.3 <u>n</u>g/µL; median: 6.19 <u>n</u>g/µL). MPXV DNA was analyzed using the broad-range FlexStar Monkeypox Virus PCR Detection Mix 1.5 (altona Diagnostics), with 21% of samples testing positive (6/28) across three sampling locations. CrAssphage-based quality control of nucleic acid extraction from torpedo sampler filters was not available, limiting assessment of sample representativeness.

Clinical mpox data were available nationally from March 2025 to February 2026, corresponding to the first reported case in Tanzania (Figure 4) [38]. Municipality-level data were available only for Tanga (March–October 2025) (Figure 5), with monthly counts exceeding five cases in March and July. The national peak occurred in March 2025 (weekly total cases ∼20), coinciding with MPXV-positive wastewater detections at two of four sampling sites. Additional wastewater detections occurred in April, June, and July 2025. No clear association was observed between estimated viral load in wastewater samples and reported clinical case numbers at national level.

**Figure 4.**
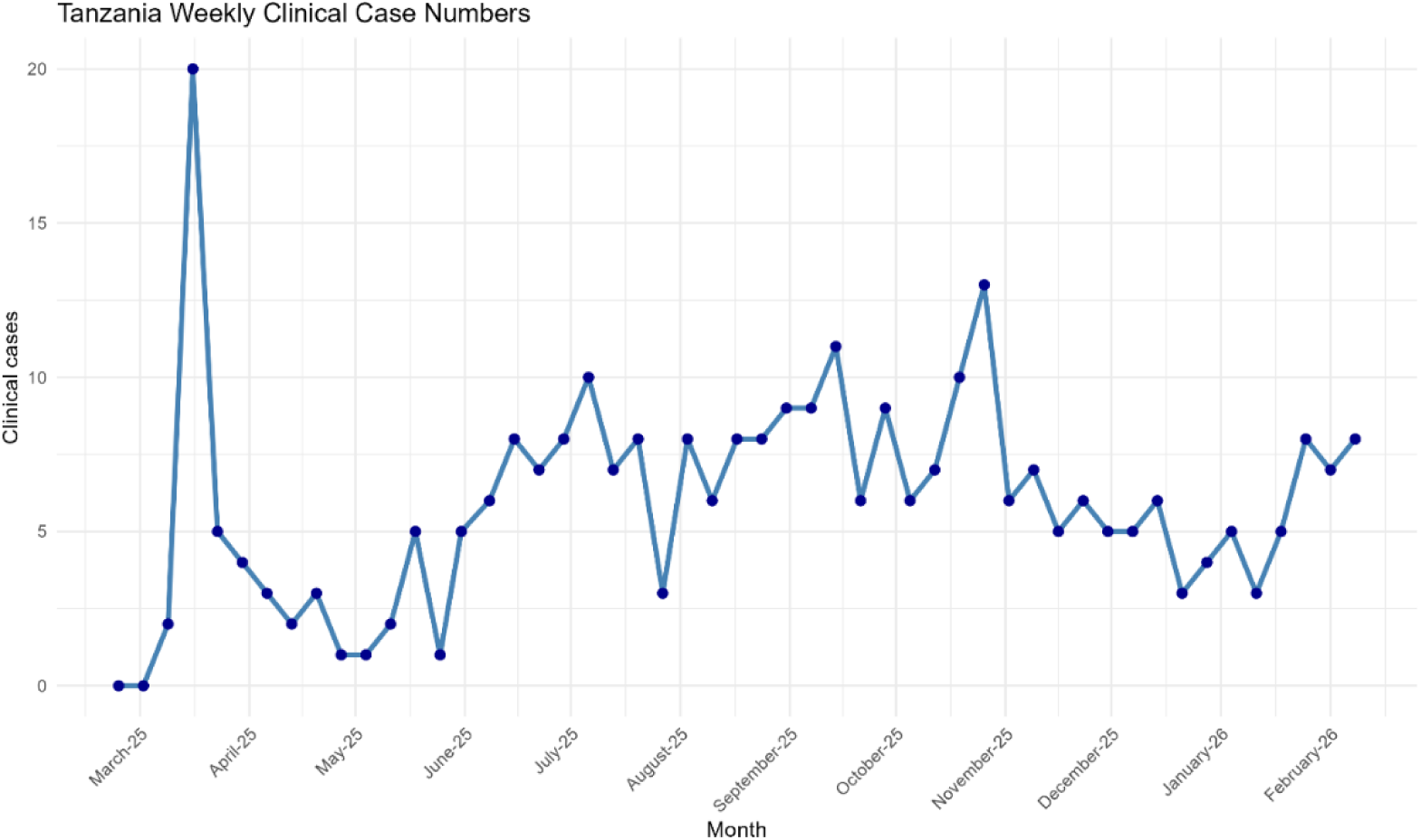
National trend of confirmed mpox clinical cases in Tanzania, March 2025- February 2026 [38]. Note: Each point represents weekly national case counts.

**Figure 5.**
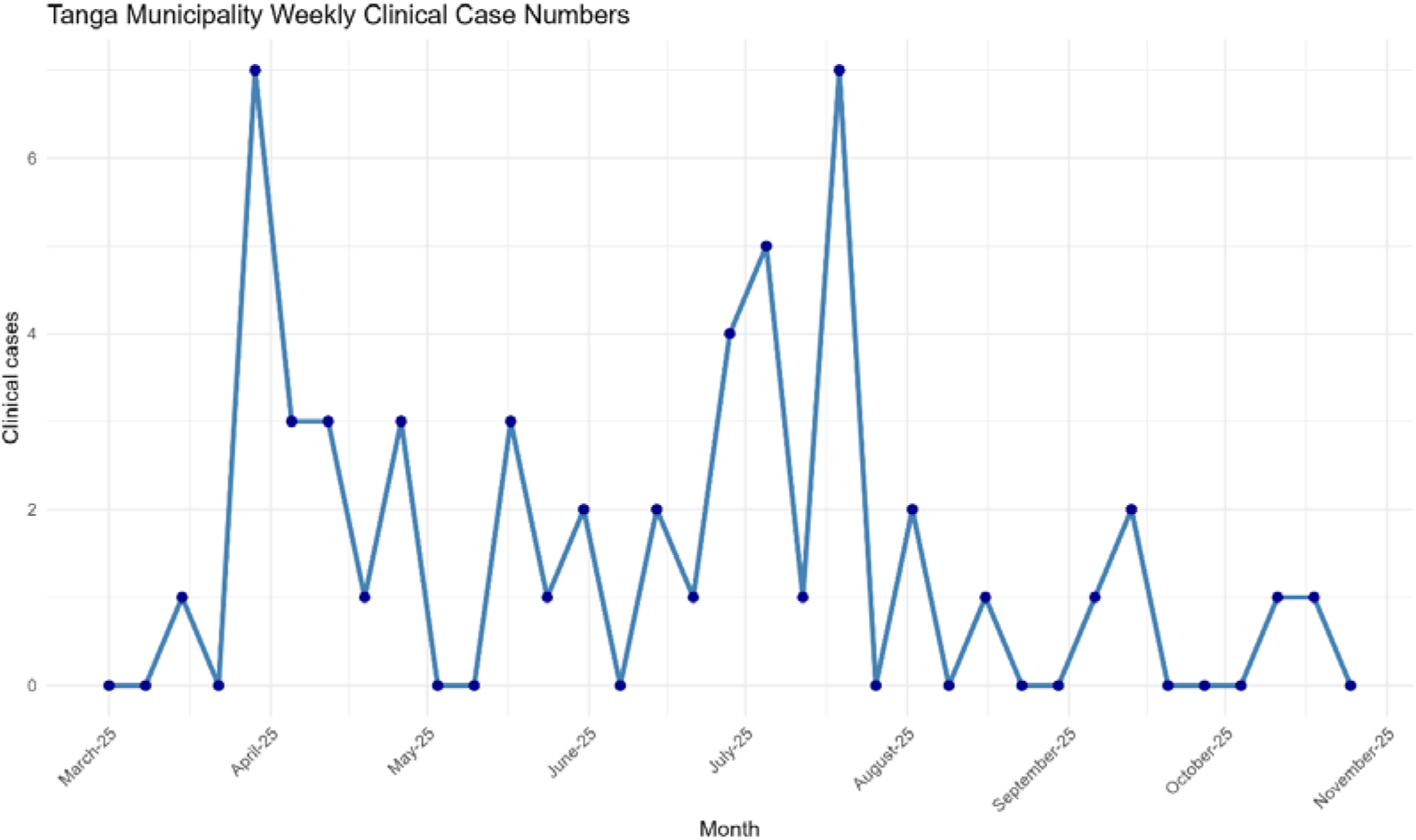
Confirmed mpox cases in Tanga region (Tanzania), March- October 2025. Data from the Tanga Regional Epidemiological Surveillance Authority. Each point represents weekly reported cases.

### 3.3 Burkina Faso

In Burkina Faso, a total of 42 passive wastewater samples were collected from four sampling locations between January 2025 and March 2026. Extracted nucleic acid concentrations ranged from 0.003 to 24.7 <u>n</u>g/µL (mean: 3.83 <u>ng</u>/µL; median: 0.39 <u>n</u>g/µL). *E. coli* enumeration indicated fecal contamination in nearly all samples (40/42). MPXV DNA and CrAssphage analyses were not completed by the end of May 2026 due to delays in processing. No confirmed mpox cases have been reported in the country to date.

## 4. Discussion

A key outcome of the ODIN-Mpox project was the successful demonstration of WWS, using torpedo passive samplers, as a functional and informative tool for MPXV detection in real-world settings. To our knowledge, torpedo passive samplers have not previously been applied in mpox WWS. MPXV DNA was consistently detected in wastewater samples from three sites in Tanzania, coinciding with the initial reporting of clinical cases, highlighting the value of environmental monitoring as a complementary indicator of community transmission. Previous studies have reported MPXV DNA in wastewater during outbreaks, supporting the interpretation that wastewater signals can reflect underlying transmission dynamics [10,42]. The detection of MPXV DNA in wastewater in Tanzania during the study period further supports the feasibility of wastewater-based mpox surveillance as a complementary tool, particularly in resource-limited settings.

In contrast, MPXV DNA was not detected in samples from the DRC, despite a high national case burden during the same period. The reason for this discrepancy remains unclear but may relate to the representativeness of the selected sampling locations, as three of four sites were surface water bodies and only one was a wastewater sampling site. In addition, reported case numbers in the sampled municipalities were low. Consequently, local transmission levels may have been below wastewater detection thresholds [20]. The interpretability of WWS results from the DRC was further limited by non-detection of fecal indicator organisms (*E. coli* and crAssphage) in majority of samples, suggesting that the collected samples did not reflect fecal inputs from the community and were not associated with viral shedding.

Of note, negative results for both *E. coli* and CrAssphage in some of the samples tested may reflect true absence of fecal contamination but may also arise from false negatives when only very small volumes of water pass through the filters or when microbial indicators are highly diluted, or if wastewater contains oxidants or other disinfecting agents [35,37,43]. Drainage canals often experience rapid flow and substantial dilution from surface water inputs, which could reduce fecal markers below detection limits [44]. Under such conditions, insufficient biomass accumulation on passive samplers can prevent reliable detection when only a low level of human-derived or fecal excreta is present.

The ODIN-Mpox project made substantial progress in laboratory readiness, surveillance capacity, and cross-institutional collaboration across African and European partner laboratories. A key achievement was the exchange of knowledge on analytical workflows to laboratories with diverse baseline capacities. Through hands-on training, protocol harmonization, and continuous technical support, the project strengthened local capacity in SSA for mpox WWS and future outbreak responses. This study focused on strengthening the operational foundation of WWS of mpox across SSA. By generating data across diverse environmental, epidemiological, and infrastructural contexts, it provided an opportunity to evaluate the feasibility, sensitivity, and potential role of wastewater monitoring as a complementary component of integrated outbreak surveillance systems. The capacity-building activities described herein may facilitate the integration of WWS into routine public health practice and support the development of early warning systems for emerging and re-emerging infectious diseases in the future. Collectively, these efforts highlight the potential of WWS as a scalable and context-sensitive approach that can complement existing preparedness and response strategies for multiple infectious disease threats across the region.

Participating laboratories in SSA demonstrated resilience by adapting workflows, sharing materials, and identifying alternative solutions despite administrative and regulatory challenges, including delays in sampling permissions and access to clinical data. These challenges varied across countries and regions and were influenced by local regulatory frameworks, operational practices, and, in some cases, broader contextual factors. Differences in laboratory capacity and wastewater system characteristics across sites required flexible implementation of workflows, as full standardization was not always feasible. While complete harmonization was not achieved, a shared methodological framework was established, improving comparability across institutions.

Future monitoring programs should prioritize careful selection of sampling sites based on accessibility, population stability, and confirmed year-round wastewater availability to ensure consistent sampling and data generation. In parallel, protocols and SOPs should be further harmonized, with emphasis on reducing turnaround times from sampling to reporting to ensure timely data availability. Continued efforts to strengthen local laboratory capacity, enhance analytical capability, and improve comparability across participating sites remain essential. Future studies should also expand the temporal and geographical scope of surveillance, increase sampling coverage and frequency, and integrate more comprehensive clinical and epidemiological datasets.

## 5. Conclusion

Overall, the ODIN-Mpox project demonstrated that WWS for mpox is feasible and informative across diverse SSA settings. The detection of MPXV signals in wastewater during periods of epidemic activity reflects both the applicability of WWS in these contexts and the increasing analytical capacity in the region. The ability to detect MPXV DNA across multiple sites, despite heterogeneous environmental and infrastructural conditions, demonstrates the operational readiness of participating institutions to contribute to integrated outbreak surveillance.

In parallel, the project strengthened local laboratory and surveillance capacity, enhanced regional collaboration, and supported the development of WWS as a scalable and sustainable component of integrated public health systems for mpox and other emerging infectious diseases. Through targeted training, protocol exchange, and sustained technical support, participating laboratories improved WWS workflows and established a durable foundation for environmental surveillance beyond the project period.

These findings contribute to the growing evidence supporting environmental surveillance for outbreak detection and situational awareness, particularly in settings where clinical surveillance may be limited or delayed. Despite infrastructural, regulatory, and capacity-related constraints, strong local engagement enabled successful implementation and provided important insights into the integration of environmental and clinical surveillance in resource-limited settings.

## Author contributions

Conceptualization: TP, RL, AK, MCT, VB, VM Validation: ESg, AT, MK, EL, PL

Formal analysis: ESg, AT, MK, EL, PL

Investigation: ESg, AT, MK, EL, PL, DS, EN, JC, GZ, EM, BK, SM, SY, MC, TT, AR, TM, TP

Writing - Original Draft: ESg, AT

Writing - Review & Editing: ESg, AT, MK, EL, PL, DS, EN, JC, GZ, EM, BK, SCM, SY, MC, TT, ESa, AR, RM, RL, AK, MCT, VB, VM, TP

Visualization: ESg

Supervision: TP, VM, VB, MCT, AK, RL

Project administration: TP, VM, VB, MCT, AK, RL Funding acquisition: TP, VM, VB, MCT, AK, RL (ESg = Elena Sgarabotto; ESa = Elisa Salmivirta)

## Data Availability

All data produced in the present work are contained in the manuscript

## References

[1] A. Gessain, E. Nakoune, Y. Yazdanpanah, Monkeypox, New England Journal of Medicine 387 (2022) 1783–1793. 10.1056/NEJMRA2208860.

[2] S. Srivastava, D. Sharma, S.B. Sridhar, S. Kumar, G.S.N.K. Rao, R.R. Budha, M.R. Babu, R. Sahu, S. Sah, R. Mehta, N.A. Giraldo-Corrales, J. Feehan, V. Apostolopoulos, A.J. Rodriguez-Morales, Comparative analysis of Mpox clades: epidemiology, transmission dynamics, and detection strategies, BMC Infect. Dis. 25 (2025) 1290-. 10.1186/S12879-025-11784-8.

3. WHO (2026). Broader transmission of mpox due to clade Ib MPXV – Global situation, (2026). https://www.who.int/emergencies/disease-outbreak-news/item/2025-DON587 (accessed June 8, 2026). World Health Organization (WHO), Geneva, Switzerland.

4. [4] WHO (2024). WHO Director-General declares mpox outbreak a public health emergency of international concern, (2024). https://www.who.int/news/item/14-08-2024-who-director-general-declares-mpox-outbreak-a-public-health-emergency-of-international-concern (accessed May 7, 2026). World Health Organization (WHO), Geneva, Switzerland.

[5] M. Kozlov, Growing mpox outbreak prompts WHO to declare global health emergency, Nature 632 (2024) 718–719. 10.1038/D41586-024-02607-Y.

6. [6] WHO (2022). WHO Director-General declares the ongoing monkeypox outbreak a Public Health Emergency of International Concern, (2022). https://www.who.int/europe/news/item/23-07-2022-who-director-general-declares-the-ongoing-monkeypox-outbreak-a-public-health-event-of-international-concern (accessed May 7, 2026). World Health Organization (WHO), Geneva, Switzerland.

[7] N. Ndembi, M. Oluwatoyin Folayan, N. Ngongo, F. Ntoumi, D. Ogoina, M. El Rabbat, J.-M. Okwo-Bele, J. Kaseya, Comment Mpox outbreaks in Africa constitute a public health emergency of continental security, Lancet Glob. Health (2024). 10.1016/S2214-109X(24)00363-2.

8. [8] DRC (2026). Déclaration officielle de la fin de l’épidémie de Mpox en République Démocratique du Congo, (2026). https://sante.gouv.cd/actualites/declaration-officielle-de-la-fin-de-lepidemie-de-mpox-en-republique-democratique-du-congo (accessed June 8, 2026).

[9] A. Tiwari, T. Kalonji, T. Miller, T. Van Den Bossche, A. Krolicka, H. Muhindo-Mavoko, P. Mitashi, M.C. Tahita, R. Lood, T. Pitkänen, V. Maketa, Emergence and Global Spread of Mpox Clade Ib: Challenges and the Role of Wastewater and Environmental Surveillance, J. Infect. Dis. 231 (2025) e825–e829. 10.1093/INFDIS/JIAF006.

[10] M.A. Islam, R. Kumar, P. Sharma, S. Zhang, P. Bhattacharya, A. Tiwari, Wastewater-Based Surveillance of Mpox (Monkeypox): An Early Surveillance Tool for Detecting Hotspots, Curr. Pollut. Rep. 10 (2024) 312–325. 10.1007/S40726-024-00299-6/TABLES/2.

11. [11] EU/EEA (2025). Surveillance of Mpox in the EU/EEA, monthly report, (2025). https://www.ecdc.europa.eu/en/infectious-disease-topics/mpox/surveillance-and-updates/surveillance-mpox-eueea-monthly-report (accessed June 8, 2026).

12. [12] WHO (2024). Surveillance, case investigation and contact tracing for mpox: interim guidance, 27 November 2024, (2024). https://www.who.int/publications/i/item/B09169 (accessed May 15, 2026). World Health Organization (WHO), Geneva, Switzerland.

[13] L.J. Beesley, D. Osthus, S.Y. Del Valle, Addressing delayed case reporting in infectious disease forecast modeling, PLoS Comput. Biol. 18 (2022) e1010115. 10.1371/JOURNAL.PCBI.1010115.

[14] S. Meschi, F. Colavita, F. Carletti, V. Mazzotta, G. Matusali, E. Specchiarello, T. Ascoli Bartoli, A. Mondi, C. Minosse, M.L. Giancola, C. Pinnetti, M.B. Valli, D. Lapa, K. Mizzoni, D.J. Sullivan, J. Ou, D. Focosi, E. Girardi, E. Nicastri, A. Antinori, F. Maggi, MPXV DNA kinetics in bloodstream and other body fluids samples, Sci. Rep. 14 (2024). 10.1038/S41598-024-63044-5.

[15] I. Brosius, C. Van Dijck, J. Coppens, L. Vandenhove, E. Bangwen, F. Vanroye, J. Verschueren, S. Zange, J. Bugert, J. Michiels, E. Bottieau, P. Soentjens, J. van Griensven, C. Kenyon, K.K. Ariën, M. Van Esbroeck, K. Vercauteren, L. Liesenborghs, Presymptomatic viral shedding in high-risk mpox contacts: A prospective cohort study, J. Med. Virol. 95 (2023). 10.1002/JMV.28769.

[16] P. Pinto, M.A. Costa, M.F.M. Gonçalves, A.G. Rodrigues, C. Lisboa, Mpox Person-to-Person Transmission—Where Have We Got So Far? A Systematic Review, Viruses 15 (2023). 10.3390/v15051074.

[17] A. Peiro-Mestres, I. Fuertes, D. Camprubi-Ferrer, M.A. Marcos, A. Vilella, M. Navarro, L. Rodriguez-Elena, J. Riera, A. Catala, M.J. Martinez, J.L. Blanco, Frequent detection of monkeypox virus DNA in saliva, semen, and other clinical samples from 12 patients, Barcelona, Spain, May to June 2022, Euro Surveill. 27 (2022). 10.2807/1560-7917.ES.2022.27.28.2200503.

[18] C. Bagutti, M.A. Hug, P. Heim, E.I. Hampe, P. Hübner, T.R. Julian, K.N. Koch, K. Grosheintz, M. Kraus, C. Schaubhut, R. Tarnutzer, E. Würfel, S. Fuchs, S. Tschudin-Sutter, Association between the number of symptomatic mpox cases and the detection of mpox virus DNA in wastewater in Switzerland: an observational surveillance study, Swiss Med. Wkly. 154 (2024) 3706–3706. 10.57187/S.3706.

[19] C. Adams, A.E. Kirby, M. Bias, A. Riser, K.K. Wong, J.W. Mercante, H. Reese, Detecting Mpox Cases Through Wastewater Surveillance - United States, August 2022-May 2023, MMWR Morb. Mortal. Wkly. Rep. 73 (2024) 37–43. 10.15585/MMWR.MM7302A3.

[20] T.R. Julian, A.J. Devaux, L. Brülisauer, S. Conforti, J.C. Rusch, C. Gan, C. Bagutti, T. Stadler, T. Kohn, C. Ort, Monitoring an Emergent Pathogen at Low Incidence in Wastewater Using qPCR: Mpox in Switzerland, Food Environ. Virol. 16 (2024) 269. 10.1007/S12560-024-09603-5.

[21] J. Oghuan, C. Chavarria, S.R. Vanderwal, A. Gitter, A.A. Ojaruega, C. Monserrat, C.X. Bauer, E.L. Brown, S.J. Cregeen, J. Deegan, B.M. Hanson, M. Tisza, H.I. Ocaranza, J. Balliew, A.W. Maresso, J. Rios, E. Boerwinkle, K.D. Mena, F. Wu, Wastewater analysis of Mpox virus in a city with low prevalence of Mpox disease: an environmental surveillance study, Lancet Regional Health. Americas 28 (2023). 10.1016/J.LANA.2023.100639.

[22] J. Rožanec, N. Kranjec, I. Obid, A. Steyer, T. Cerar Kišek, T. Koritnik, M. Fafangel, A. Galičič, Wastewater Surveillance of Mpox during the Summer Season of 2023 in Slovenia, Infect. Dis. Rep. 16 (2024) 836–845. 10.3390/IDR16050065.

[23] O.E. Hart, R.U. Halden, Computational analysis of SARS-CoV-2/COVID-19 surveillance by wastewater-based epidemiology locally and globally: Feasibility, economy, opportunities and challenges, Science of The Total Environment 730 (2020) 138875. 10.1016/J.SCITOTENV.2020.138875.

24. [24] J. Kinyua, N. Negreira, A. McCall, T. Boogaerts, C. Ort, A. Covaci, A. Nuijs, The emerging field of wastewater-based epidemiology (WBE), (2019).

[25] C.S. Truyens, D.M. Berendes, M.E. Cantrell, A.L. Kossik, K.M. McCarthy, A.S. Mehrotra, J.L. Murphy, S. Pillay, S.J. Raj, M.S. Ramaswamy, H. Yakubu, R.H. Holm, Advancing wastewater and environmental surveillance in LMICs for public health response and SDG data gaps, Npj Clean Water 2025 8:1 8 (2025) 95-. 10.1038/s41545-025-00523-w.

[26] B. Baker, J.A. Baz Lomba, J. Bitilinyu-Bangoh, A. Berglöf, S. Bombaywala, T. Calvert-Joshua, B. Kaboré, P. Kingpriest, T. Lang, J.I. Levy, P. Lompo, E. Lyimo, L. Martens, H.M. Mavoko, B. Mesuere, N. Moremi, N. Mulder, S.L. Ndure, M.A. Rameto, T.F. Rinke de Wit, H. Sebukoto, E. Smith, M.C. Tahita, V.M. Tevuzula, B.A. Tippett Barr, A. Tiwari, T. Tran, E. Ubomba-Jaswa, T. Van Den Bossche, D. Wolday, A. Krolicka, V. Baraka, T. Pitkänen, R. Lood, Project ODIN: advancing environmental genomic surveillance for public health across sub-Saharan Africa, Lancet Microbe (2026). 10.1016/J.LANMIC.2026.101426.

[27] T. Miller, K. Valkama, V. Baraka, M.C. Tahita, V. Maketa, E. Lyimo, H. Sebukoto, P. Lompo, B. Kaboré, Z. Garba, S. Yougbare, M. Kabena, E. Ngelesi, J. Clever, P. Mkama, M. Chuwa, S. Mnyawonga, O. Nyholm, A.M. de R. Husman, T. Pitkänen, Towards a Framework for the Successful Implementation of Wastewater and Environmental Surveillance for Diarrhoeal Diseases in Sub-Saharan Africa, (2026). 10.20944/PREPRINTS202602.1105.V1.

[28] H. Al-Hello, O. Nyholm, T. Pitkänen, E. Salmivirta, A. Vainio, K. Valkama, V. Baraka, E. Lyimo, S. Mnyawonga, E. Msoma, H. Sebukoto, M. Chuwa, J. Claver, I. Mauki, N. Moremi, Z. Garba, P. Lompo, Y. Sibidou, M. Tahita, M. Kabena Kabengele, V. Maketa, E. Ngelesi, M. Kisala, B. Modadra Madakpa, T. Bodisa Matamu, M.-A. Kavira Muhindo, M. Ngimba, P. Kabuyaya Nzoloka, E. Tsiwedi Tsilabia, ODIN Laboratory Handbook Standard Operating Procedures for Pre-treatment of Environmental Samples, Pathogen Analytics and Whole Genome Sequencing, ODIN Wastewater Surveillance Project (2025). 10.48060/TGHN.154.

[29] T. Miller, T. Pitkänen, O. Nyholm, K. Valkama, M. C. Tahita, V. Baraka, V. Maketa, E. Lyimo, H. Sebukoto, P. Lompo, M. Kabena, A. Krolicka. ODIN Protocol for Environmental Pathogen Surveillance in Sub-Saharan Countries. 10.48060/tghn.155.

[30] C. Schang, N.D. Crosbie, M. Nolan, R. Poon, M. Wang, A. Jex, N. John, L. Baker, P. Scales, J. Schmidt, B.R. Thorley, K. Hill, A. Zamyadi, C.W. Tseng, R. Henry, P. Kolotelo, J. Langeveld, R. Schilperoort, B. Shi, S. Einsiedel, M. Thomas, J. Black, S. Wilson, D.T. McCarthy, Passive Sampling of SARS-CoV-2 for Wastewater Surveillance, Environ. Sci. Technol. 55 (2021) 10432–10441. 10.1021/ACS.EST.1C01530.

[31] Y. Li, V.A. Olson, T. Laue, M.T. Laker, I.K. Damon, Detection of monkeypox virus with real-time PCR assays, Journal of Clinical Virology 36 (2006) 194–203. 10.1016/J.JCV.2006.03.012.

[32] Y. Li, H. Zhao, K. Wilkins, C. Hughes, I.K. Damon, Real-time PCR assays for the specific detection of monkeypox virus West African and Congo Basin strain DNA, J. Virol. Methods 169 (2010) 223–227. 10.1016/J.JVIROMET.2010.07.012.

[33] L. Schuele, L.M. Masirika, J.C. Udahemuka, F.B. Siangoli, J.B. Mbiribindi, P. Ndishimye, F.M. Aarestrup, M. Koopmans, B.B. Oude Munnink, R. Molenkamp, M. Boter, H. Cassidy, L.M. Mambo, G. Overbeek, J.P. Musabyimana, J. Ndoli, S. Otani, Real-time PCR assay to detect the novel Clade Ib monkeypox virus, September 2023 to May 2024, Eurosurveillance 29 (2024) 2400486. 10.2807/1560-7917.ES.2024.29.32.2400486/CITE/REFWORKS.

[34] M.L. Devane, E. Moriarty, L. Weaver, A. Cookson, B. Gilpin, Fecal indicator bacteria from environmental sources; strategies for identification to improve water quality monitoring, Water Res. 185 (2020) 116204. 10.1016/J.WATRES.2020.116204.

[35] E. Stachler, C. Kelty, M. Sivaganesan, X. Li, K. Bibby, O.C. Shanks, Quantitative CrAssphage PCR Assays for Human Fecal Pollution Measurement, Environ. Sci. Technol. 51 (2017) 9146–9154. 10.1021/ACS.EST.7B02703/SUPPL_FILE/ES7B02703_SI_001.PDF.

[36] J. Langeveld, R. Schilperoort, L. Heijnen, G. Elsinga, C.E.M. Schapendonk, E. Fanoy, E.I.T. de Schepper, M.P.G. Koopmans, M. de Graaf, G. Medema, Normalisation of SARS-CoV-2 concentrations in wastewater: The use of flow, electrical conductivity and crAssphage, Science of The Total Environment 865 (2023) 161196. 10.1016/J.SCITOTENV.2022.161196.

[37] K. Farkas, E.M. Adriaenssens, D.I. Walker, J.E. McDonald, S.K. Malham, D.L. Jones, Critical Evaluation of CrAssphage as a Molecular Marker for Human-Derived Wastewater Contamination in the Aquatic Environment, Food Environ. Virol. 11 (2019) 113. 10.1007/S12560-019-09369-1.

38. [38] WHO (2026). Global Mpox Trends. https://worldhealthorg.shinyapps.io/mpx_global/#sec-clades (accessed May 26, 2026). World Health Organization (WHO), Geneva, Switzerland.

39. [39] WHO (2026). Mpox: Multi-country External Situation Report no. 65. 30 April 2026, Emergency situation update. World Health Organization (WHO), Geneva, Switzerland.

40. [40] Article: Sampling begins across ODIN sites • ODIN Wastewater Surveillance Project, (n.d.). https://odin-wsp.tghn.org/articles/pilot-sampling-across-odin-partner-countries/ (accessed May 26, 2026).

41. [41] Article: ODIN Wastewater Surveillance Project: Sampling campaign completed • ODIN Wastewater Surveillance Project, (n.d.). https://odin-wsp.tghn.org/articles/odin-wastewater-surveillance-project-sampling-campaign-completed/ (accessed May 26, 2026).

[42] K. Brighton, S. Fisch, H. Wu, K. Vigil, T.G. Aw, Targeted community wastewater surveillance for SARS-CoV-2 and Mpox virus during a festival mass-gathering event, Science of The Total Environment 906 (2024) 167443. 10.1016/J.SCITOTENV.2023.167443.

[43] M. Geissler, R. Mayer, B. Helm, R. Dumke, Food and Environmental Virology: Use of Passive Sampling to Characterize the Presence of SARS-CoV-2 and Other Viruses in Wastewater, Food Environ. Virol. 16 (2024) 25–37. 10.1007/S12560-023-09572-1.

[44] J.G. Kimaro, V. Scharsich, A. Kolb, B. Huwe, C. Bogner, Distribution of traditional irrigation canals and their discharge dynamics at the southern slopes of Mount Kilimanjaro, Front. Environ. Sci. 7 (2019) 423701. 10.3389/FENVS.2019.00024/TEXT.

